# Mobility patterns, activity locations, and tuberculosis in Nairobi, Kenya

**DOI:** 10.1101/2024.09.12.24313589

**Authors:** Khai Hoan Tram, Jane Ong’ang’o, Richard Kiplimo, Thomas R. Hawn, Videlis Nduba, David J. Horne, Jennifer M. Ross

**Affiliations:** Division of Allergy and Infectious Diseases, University of Washington, Seattle, WA, USA; Centre for Respiratory Diseases Research, Kenya Medical Research Institute, Nairobi, Kenya; Amref Health Africa, Nairobi, Kenya; Division of Pulmonary, Critical Care, and Sleep Medicine, University of Washington, Seattle, WA, USA

## Abstract

**Background:** Annually, over 3 million people develop TB but are not diagnosed and treated. We aimed to characterize the mobility patterns and activity locations of people with TB in an urban, high-burden setting to inform future active case finding (ACF) efforts.

**Methods:** We conducted a population-based TB prevalence survey in Nairobi, Kenya, in 2022. Participants aged ≥15 years with TB symptoms or a suggestive chest x-ray submitted sputum for Xpert Ultra and culture. We collected data on individual activity locations and mobility and evaluated their association with the risk of pulmonary TB.

**Results:** The prevalence survey enrolled 6369 participants across nine clusters. There were significant differences in mobility patterns and activity locations between sexes and age groups. Mobility factors were not significantly associated with TB. In the adjusted analysis, age group 45–54 (OR 2.45), male sex (OR 2.95), and use of a social activity location (OR 1.96) were significantly associated with a higher risk of TB.

**Conclusions:** We did not find a significant association between mobility patterns and TB but found a positive association between reported ‘social’ activity locations and TB. Identification of ‘social’ activity locations, particularly bars, provides important insight into possible venues for spatially-targeted ACF activities.

## Introduction

Tuberculosis (TB) remains one of the leading infectious causes of death globally, with an estimated 10.6 million people with incident TB and 1.3 million deaths in 2022.^1^ TB case notifications increased to 7.5 million in 2022 after several years of COVID-19 pandemic-related disruptions, still leaving over 3 million people annually who develop TB but who are not diagnosed or initiated on treatment.^1^ This case-detection gap critically impedes global efforts to end the TB epidemic as individuals with undiagnosed TB likely contribute substantially to ongoing transmission and are at higher risk for death if treatment is delayed or not initiated.^2,3^ Identification of people with TB in the community through active case finding (ACF) can help to achieve earlier diagnosis and treatment and possibly prevent onward transmission.^4^

Mobile individuals, and in particular migrants, may be at higher risk for TB and could potentially benefit from screening through ACF.^5–7^ For example, migrant mineworkers in southern Africa experience some of the highest TB incidence rates in the world due to exposure to multiple risk factors, such as HIV and poverty, and frequent health care disruptions.^8^ Mobility on a smaller scale, such as one’s daily commute to work or school, may also contribute to the TB epidemic. Several studies from Cape Town, South Africa, of CO_2_ levels and social contacts suggest public transportation as one possible site for TB transmission.^9–11^ Similarly, in Lima, Peru, the risk of TB among workers in the informal public transport sector was significantly higher than in the general population.^12,13^ Despite playing a potentially important, and varied, role in the transmission of TB, there remain large gaps in our understanding of human mobility as a risk factor for TB across different settings or even how to properly measure it.^14,15^

A second challenge for implementing ACF, after identifying a priority population for screening, is to determine where to deploy ACF activities in a community. Traditional ACF may encompass a range of activities such as door-to-door screening or community mobilization,^4,16^ though these activities can be costly when implemented over a broad area. In contrast, a spatially-targeted ACF strategy focuses on geographic areas or locales with suspected high burden of disease, with the potential benefit of needing to screen fewer people to identify people with TB.^17^ Activity spaces, which are locations or sites where an individual spends their time, present one possible solution to the question of where to geographically deploy ACF. One study mapped ‘activity spaces’ for patients with culture-positive TB residing in Shinjuku City, Japan and located the peak density of cases at the

Shinjuku Railway Station that was over 12 times greater than average.^18^ In a study in a rural Ugandan township, 6 out of 15 (40%) genotype-matched TB clusters shared potential epidemiologic links based on either a shared physical location or geographically neighboring location (within 100 meters) in the time leading up to diagnosis.^19^ Likewise, in a case-control study in Lima, Peru, activity spaces were obtained from both multidrug-resistant (MDR) TB cases and negative controls and demonstrated that person-time (in hours) in healthcare venues, school, and transportation venues was significantly associated with MDR TB diagnosis.^20^

Emerging strategies of disease control require understanding the role of human mobility and activity spaces on the TB epidemic, both at an individual and population level,^14,21^ and tailoring interventions such as active case finding to populations who are at increased risk.^22,23^ Our goal in this analysis of mobility and activity locations data, collected in conjunction with a TB prevalence survey, is to evaluate TB with regards to geographic and behavioral risk factors in an urban, high-burden setting.

## Methods

### Prevalence survey study design and methods

This analysis leverages data from a cluster-based cross-sectional TB prevalence survey that enrolled participants in Nairobi, Kenya, from May to December 2022.^24^ The survey aimed to determine the prevalence of bacteriologically-confirmed pulmonary tuberculosis among people ages 15 years and older in nine neighborhoods in Nairobi that were previously surveyed in the 2015-2016 Kenya National TB Prevalence Survey.^25^ Prevalence survey details are published elsewhere. In brief, people aged ≥ 15 years who had lived within the neighborhood boundaries for at least 30 days were invited to a nearby mobile field site where they completed questionnaires, a TB symptom screen, and digital chest X-ray. Participants with an abnormal radiograph suggestive of TB or positive symptom screen (reporting a cough of any duration) were eligible to submit 2 sputum samples (spot and morning) for testing by GeneXpert MTB/RIF Ultra (Xpert Ultra, Cepheid, Sunnyvale, CA, USA) and using Mycobacterium Growth Indicator Tubes (MGIT 960, Becton, Dickinson and Company, Franklin Lakes, NJ, USA). All participants with a positive culture or positive Xpert Ultra result (including ‘trace positive’) were referred to local health clinics for TB treatment.

In the primary analysis of the prevalence survey, a study participant was defined as having prevalent pulmonary TB if the Xpert Ultra result was positive (at a semiquantitative level that was greater than ‘trace positive’ [i.e., very low, low, medium, or high]) or at least one sputum culture was positive for *M. tuberculosis.* In contrast, in this analysis of mobility patterns and activity locations, all participants with a positive *Mtb* culture or positive Xpert Ultra result (including ‘trace positive’) were considered to be positive.

### Measurement of mobility patterns and activity locations

Several components of geographic mobility were collected via a tablet-based questionnaire administered by a trained research assistant. First, dimensions of mobility relating to daily commute, specifically commuting to work, were obtained from the subset of participants who reported a work location. These data include mode of transportation (e.g. walk, bus, taxi, etc.), distance of commute (in kilometers), length of commute (in hours), expense (in Kenyan shillings), and use of transit hubs. Second, participants were asked about overnight trips in the past month, including frequency, duration, and reason for travel. Third, participants were asked about changes in residence over the past 5 years. Finally, data on activity locations were collected, as participants were asked to list the top four places where they spent their waking hours, as well as the top four places they visited in the community on weekdays and on weekends. Participants could list up to 12 possible locations (though many places were repeated). The question structure did not explicitly ask for participants to rank these places by the amount of time they spent there. The activity locations data were collected as free text responses and manually categorized into one of six categories: home, work, school, market, social (e.g. bars, gaming, sports club, social halls), or church.

### Statistical analysis

We first generated descriptive summary statistics for mobility patterns and activity locations for the total population and stratified by sex (female & male) and age group (15-24, 25-34, 45-54, 55-64, and 65+). Comparisons between groups were performed using chi-square test. We then used logistic regression to evaluate the association between demographic characteristics, mobility patterns, ‘mobility’ class membership, and reported activity locations with our primary outcome of prevalent pulmonary TB, i.e., odds of being a TB case. The adjusted analysis included age and sex. A secondary analysis explored these same independent variables with odds of positive symptom screen (e.g. cough of any duration, fever, night sweats, or weight loss).

In the multivariate analysis, reporting a particular category of activity location does not necessarily mean that that activity location category was the first reported place. The activity location exposure variables indicate any reporting of a particular activity location, whether in the top 4 places where people spent their waking hours, the top 4 places visited on weekdays, or the top 4 places visited on weekends.

Given the complexity and multidimensionality of human mobility, we performed latent class analysis (LCA) to construct one latent variable from six of our observed variables capturing various features of mobility (**Table S1**). LCA is a statistical technique used to identify different subgroups or ‘latent classes’ within a population that may share certain characteristics and can explain patterns of observed behaviors or variables.^26^ LCA assigns a probability of class membership for each individual in the dataset, and in this manner allows one to identify which individuals should be grouped together based on a chosen set of variables.^26^ Since we are interested in mobility as a particular mode of behavior and have measured various components of mobility (e.g. daily commute, overnight trips, changes in residence), LCA can also help us to identify different ‘mobility’ subgroups or profiles within our population.^14^ Models with between 1-4 latent classes were tested, and we selected the best fitting latent class model based on the Akaike information criterion (AIC) and Bayesian information criterion (BIC).

### Ethics statement

This study was approved by the KEMRI Scientific and Ethics Review Unit (KEMRI/SERU/3988) and the University of Washington Institutional Review Board (UW STUDY00009209). Risks and benefits of participating in the study were explained to each participant. Each participant signed a written informed consent form. For those participants under the age of 18, guardian consent and participant assent were obtained.

## Results

### Study population

The 2022 Nairobi Prevalence Survey enrolled a total of 6369 participants (83.3% enrollment out of 7644 eligible). Of those enrolled, 1504 participants (23.6%) completed the work commute questionnaire and 5748 (90.2%) completed questions regarding activity locations. Over 99.5% of participants answered questions about overnight trips away from home in the past month and changes in residence over the past 5 years. Only those participants who reported that they work outside of the home or attend school were administered the work commute questions. Compared with other enrolled participants, the work commute survey subgroup was significantly younger, had a greater percent that were male, and were more likely to be employed in the private sector or be a student (**Table 1**).

**Table 1.** Sociodemographic characteristics of participants.

|  | Overall Prevalence<br>survey<br>N = 6369 enrolled | Mobility patterns |  | Activity<br>locations |  |
| --- | --- | --- | --- | --- | --- |
|  |  | n = 1504 (23.6%) | p-value* | n = 5748 (90.2%) | p-value* |
| **Age, median [IQR]** | 33 [24 – 42] | 30 [19 – 41] | **<0.001** | 33 [24 – 43] | 0.235 |
| **Age group** |  |  |  |  |  |
| 15 – 24 | 1663 (26.1%) | 568 (37.8%) | **<0.001** | 1496 (26.0%) | 0.786 |
| 24 – 34 | 1901 (29.8%) | 367 (24.4%) |  | 1706 (29.7%) |  |
| 35 – 44 | 1450 (22.8%) | 280 (18.6%) |  | 1307 (22.7%) |  |
| 45 – 54 | 751 (11.8%) | 203 (13.5%) |  | 686 (11.9%) |  |
| 55 – 64 | 393 (6.1%) | 64 (4.3%) |  | 357 (6.2%) |  |
| 65+ | 213 (3.3%) | 22 (1.5%) |  | 196 (3.4%) |  |
| **Sex** |  |  |  |  |  |
| Female | 4184 (65.7%) | 793 (52.7%) | **<0.001** | 3759 (65.4%) | 0.151 |
| Male | 2187 (34.3%) | 711 (42.3%) |  | 1989 (34.6%) |  |
| **HIV status (self-reported)** |  |  |  |  |  |
| Positive | 138 (3.3%) | 28 (2.7%) | 0.199 | 121 (3.1%) | **0.012** |
| Negative | 4016 (96.7%) | 1008 (97.3%) |  | 3745 (96.9%) |  |
| **Number of household members, mean (std)** | 2.8 (±1.7) | 3.0 (±1.7) | **<0.001** | 2.8 (±1.8) | **<0.001** |
| **Occupation** |  |  |  |  |  |
| Self-employed | 2774 (43.6%) | 591 (39.4%) | **<0.001** | 2495 (43.4%) | **0.001** |
| Employed by government | 84 (1.3%) | 57 (3.8%) |  | 80 (1.4%) |  |
| Employed in private sector | 615 (9.7%) | 375 (25.0%) |  | 560 (9.8%) |  |
| Pupil/student | 584 (9.1%) | 424 (28.2%) |  | 506 (8.8%) |  |
| Housewife | 880 (13.8%) | 6 (0.4%) |  | 799 (13.9%) |  |
| Unemployed | 1311 (20.6%) | 21 (1.4%) |  | 1205 (21.0%) |  |
| Other | 120 (1.9%) | 28 (1.9%) |  | 100 (1.7%) |  |

\* Wilcoxon rank-sum test, Chi-square tests, and t-tests comparing those enrolled participants who did answer or did not answer the mobility patterns and activity locations questions, respectively

### Work commute

Overall, the majority of the 1504 respondents to the work commute questionnaire walked to work (68.3%) or used a bus or taxi (28.8%), with only a small fraction riding boda boda (motorbike taxis) or using other means of transportation (**Table 2**). Women more likely to walk than men (73.6% vs 62.3%, p<0.001) and less frequently used the bus or taxi (24.5% vs 33.5%, p<0.001). Across age groups, bus/taxi ridership was highest among those ages 15-24 (34.7%) and lowest among those ages 45-54 (22.2%). The median commute distance was 2km [IQR 1-5]. One-third of commuting trips (32.7%) were below 1km. Men, on average, had a longer commute distance than women (mean 7.6km vs 5.2km) and longer commute time (mean 1.85 hours vs 1.32 hours). Only 241 of the total participants (3.8%) reported spending time at a transit station, with a median time of 10 minutes [IQR 5-30].

**Table 2.**
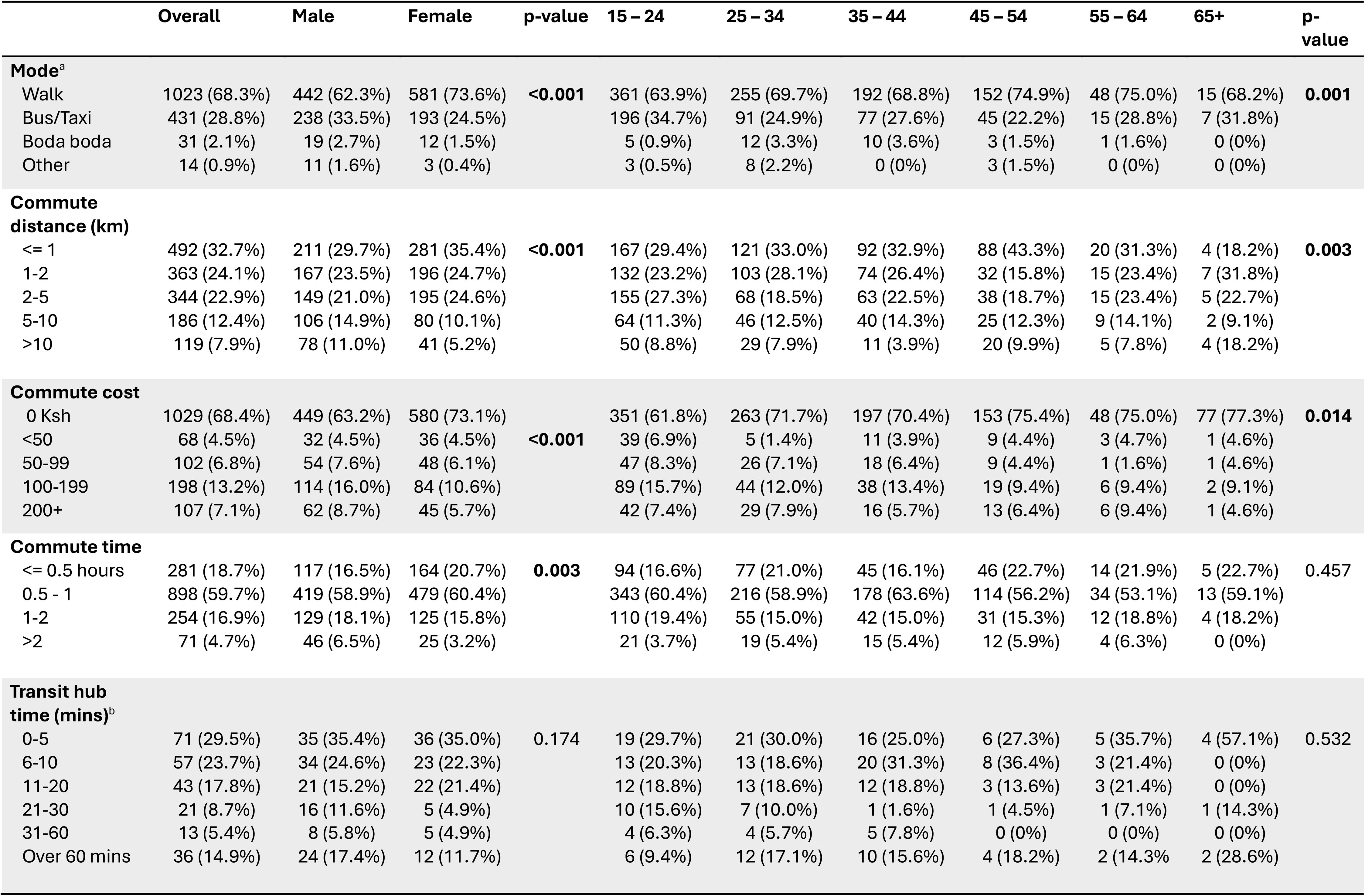
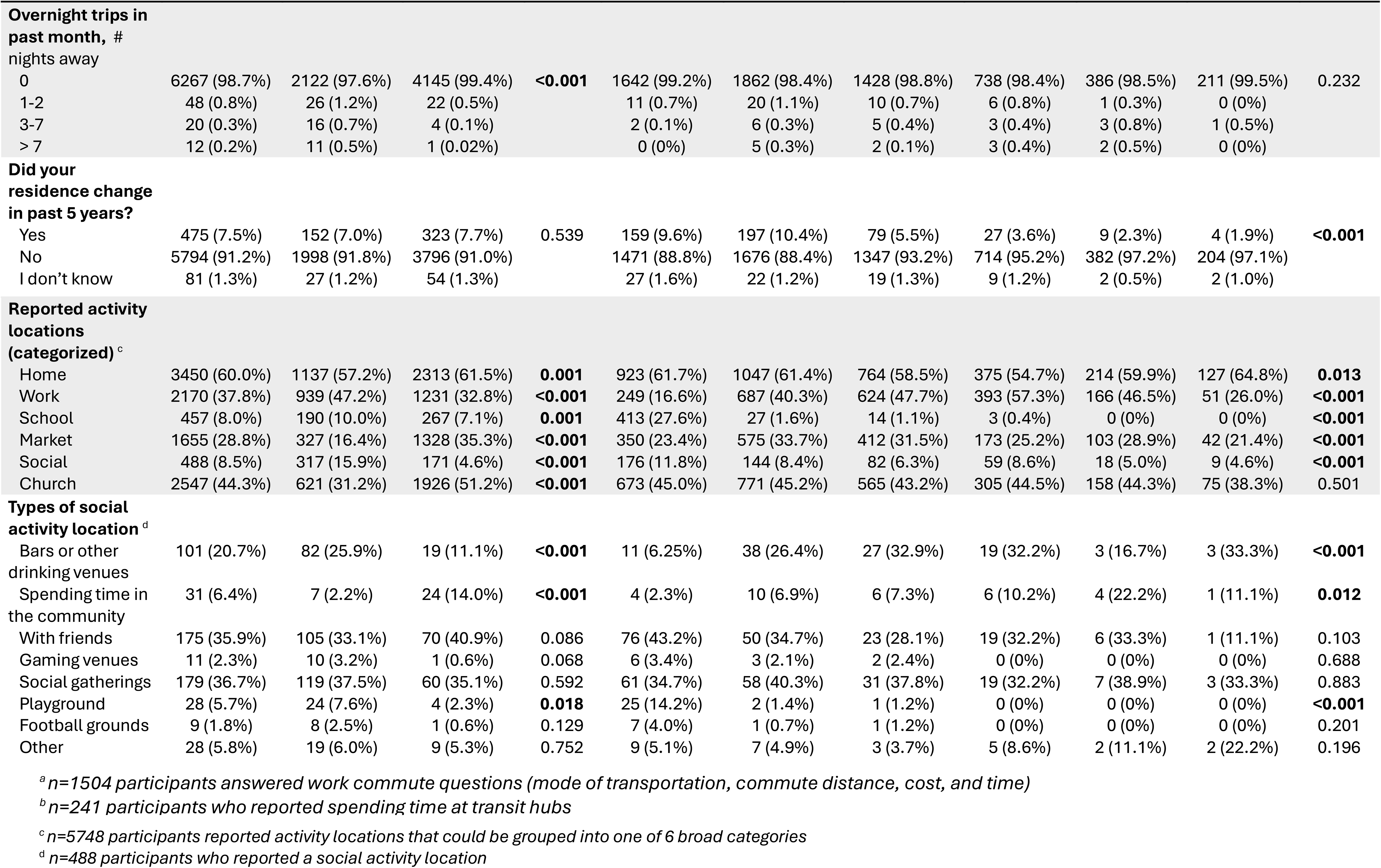
Mobility patterns and activity locations by sex and age group.

### Activity locations

A total of 5748 participants responded to the activity locations questions. In all, 24,448 responses were recorded (mean 4.3 ± 2.3 places reported per participant), of which 17,943 (73.4%) could be manually coded into one of six activity locations categories. Overall, home was the most frequently reported location, followed by church, work, and market (**Table 2**). These activity location categories differed significantly by sex—for example, women were twice as likely (35.3% vs 16.4%) to report ‘market’ and men were three times as likely (15.9% vs 4.6%) to report a ‘social’ activity locations— and age group. Reported activity locations also varied significantly across neighborhoods (p-value <0.001). **Figure 1** presents the first and second reported activity locations (of 12 maximum) by men and women in the nine neighborhoods surveyed in this study. Heterogeneity in the categories of reported activity locations is clear across these geographic clusters.

**Figure 1.**
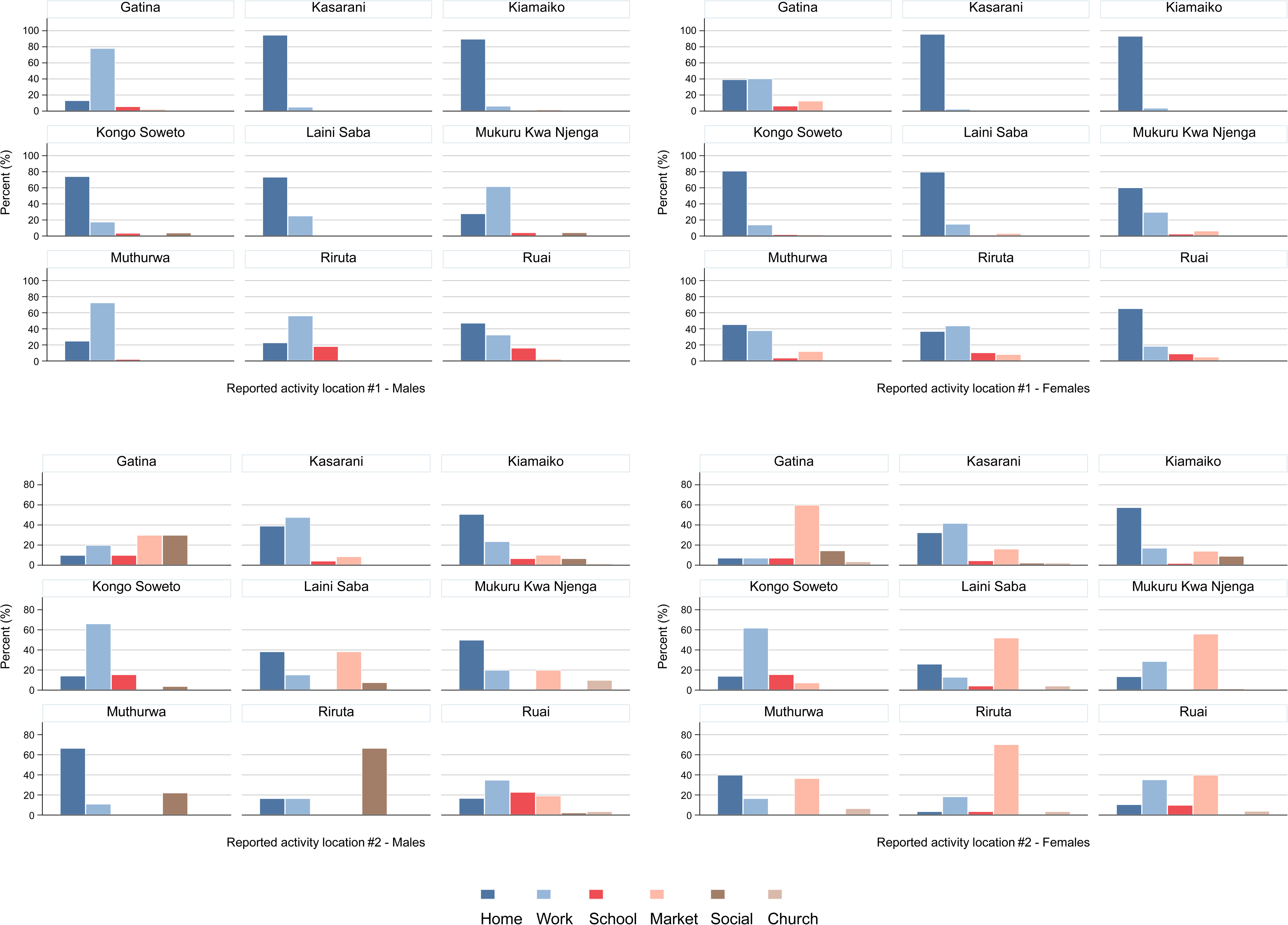
Reported activity locations by sex across 9 neighborhoods in Nairobi, Kenya. Categories of activity locations reported by men (left) and women (right). The two top charts depict the first reported activity location by participants when asked to list the top 4 places where they spent most of their waking hours in the past year, and the two bottom charts depict the second reported activity location. These data do not necessarily represent a ranked ordering in terms of time spent in these places.

Of those reporting ‘social’ activity locations (n=488), 101 (20.7%) reported bars or drinking venues, 175 (35.9%) reported being with friends, and 179 (36.7%) reported going to social places or gatherings. Other ‘social’ activity locations included spending time in the community (reported by 6.4%), playground (5.7%), gaming (2.3%), football grounds (1.8%), or other activities (5.8%).

**Figure 2** illustrates a two-mode network of shared activity location categories within a single neighborhood in the study. Each enrolled participant in the neighborhood is represented by a single blue circular node with lines connecting each individual to their reported activity location categories represented by red squares. This visualization demonstrates the types of shared spaces in a community that may be targeted for active case finding activities.

**Figure 2.**
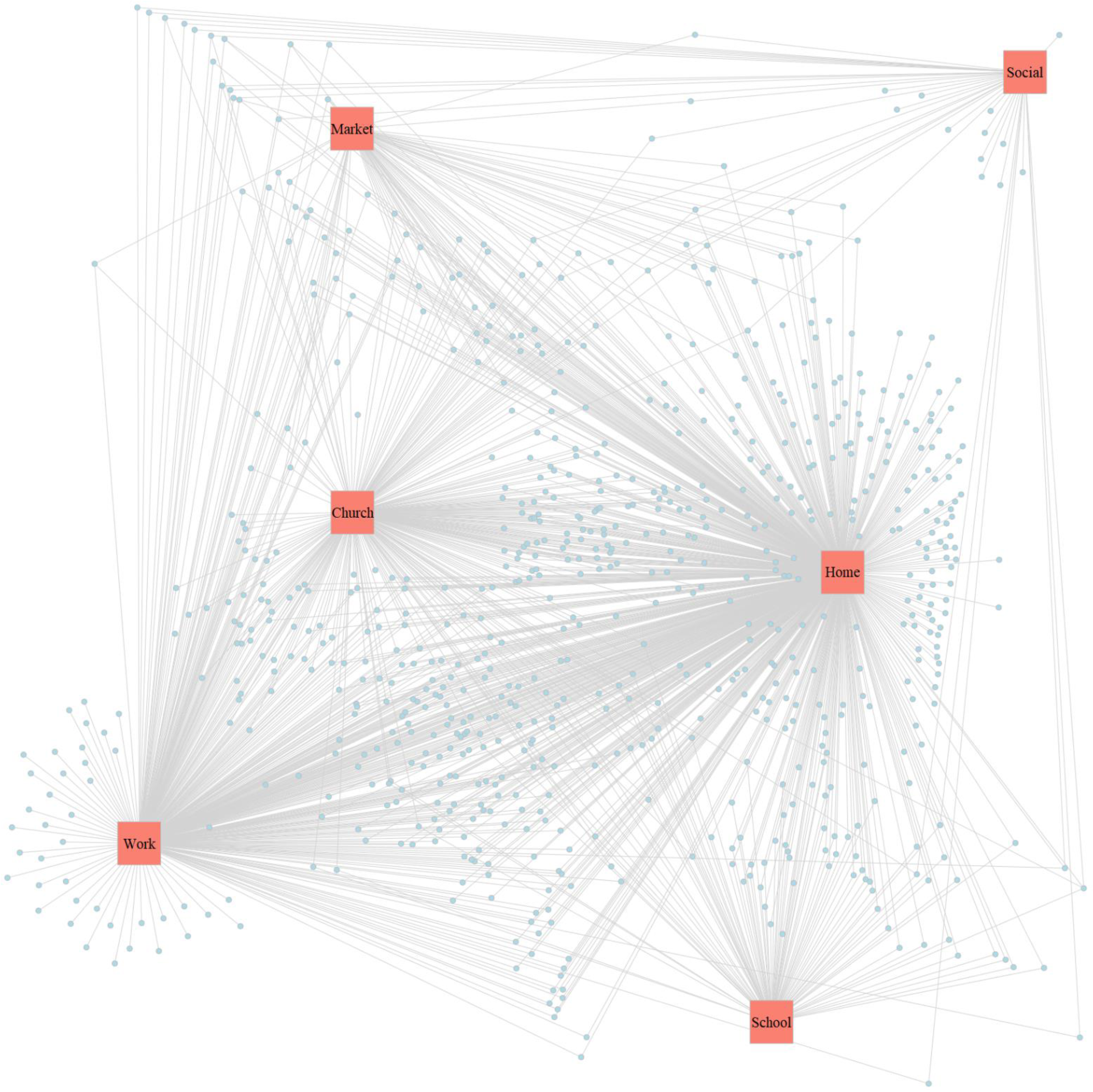
Visualization of a two-mode network with shared activity location categories. Representative bipartite network illustrating shared activity location categories within one surveyed neighborhood in Nairobi, Kenya. Blue circles represent individual participants and red squares represent categories of activity locations (e.g. home, work, school, market, church, or social). Each node (blue circle) is connected to one or more activity location categories (red squares), depending on the places that they reported.

### Latent class analysis – classification of mobility

To examine whether there were distinct patterns of mobility amongst participants, we used a latent class analysis which incorporated a range of mobility variables. A model for mobility with two latent classes was selected as it had a better fit compared to a single class based on a lower BIC and AIC (**Table S2**). Models with three and four latent classes did not converge after 300 iterations. One class was characterized as “higher mobility” and the other “lower mobility” based on the estimated marginal probabilities for six mobility indicator variables. Each participant was assigned group membership to their most likely ‘mobility’ class based on posterior probabilities. Of the 1,499 participants included in the latent class analysis—limited by the number of responses to the work commute questionnaire—45.4% were in the “higher mobility” class and 54.6% were in the “lower mobility” class. The most significant differences between the two classes, based on estimated marginal probabilities, were as follows: using the bus, taxi, or other mode of transportation besides walking (69.6% vs 3.1%), commuting distance greater than 2km (92.0% vs. 6.3%), and commute time greater than 0.5 hours (97.0% vs 69.7%). There were much smaller differences between the two classes with regard to use of transit hubs, overnight trips in the past month, or changes in residence in the past 5 years (**Table S3 & Figure S1**).

### Association of demographics, mobility, & activity locations with TB and symptom screen

Next, we examined whether demographic characteristics, mobility, or activity locations were associated with TB. In total, there were 83 enrolled participants with a diagnosis of TB (either positive Xpert Ultra including ‘trace positive’ and/or positive *Mtb* culture). Of these, 16/83 (19.3%) responded to the work commute questions and 73/83 (88.0%) reported activity locations. Out of the 83 participants with a TB diagnosis, 38 (45.8%) were Xpert Ultra ‘trace positive’ only without a positive *Mtb* culture.

In the unadjusted analysis, older age groups, male sex (OR 2.94, 95%CI 1.89, 4.58), and reporting a social activity location (OR 2.15, 95%CI 1.15, 4.02) were all significantly associated with a diagnosis of active TB in the prevalence survey (**Table 3**). Mobility factors, specifically mode of transportation, commute distance, and commute time, as well as overnight trips and changes in residence were not significantly associated with active TB diagnosis, nor was membership in the ‘higher mobility’ latent class. Adjusting for age and sex: age group 45 – 54 years (OR 2.45, 95%CI 1.05, 5.71), male sex (OR 2.95, 95%CI 1.76, 4.95), and reporting a social activity location (OR 1.96, 95%CI 1.03, 3.74) were significantly associated with higher odds of active TB diagnosis.

**Table 3.** Association of mobility & activity locations with active TB diagnosis and positive symptom screen.

|  | Bacteriologically-confirmed TB |  |  |  | Reporting any symptoms |  |  |  |
| --- | --- | --- | --- | --- | --- | --- | --- | --- |
|  | Unadjusted OR | p-value | Adjusted OR<sup>b</sup> | p-value | Unadjusted OR | p-value | Adjusted OR<sup>c</sup> | p-value |
| **Age group** (n=6371) |  |  |  |  |  |  |  |  |
| 15 – 24 | Ref |  | Ref |  | Ref |  | Ref |  |
| 24 – 34 | 2.16 (1.07, 4.36) | **0.032** | 1.84 (0.85, 3.98) | 0.122 | 0.97 (0.81, 1.17) | 0.776 | 1.00 (0.81, 1.23) | 0.984 |
| 35 – 44 | 2.31 (1.12, 4.79) | **0.024** | 1.83 (0.81, 4.14) | 0.149 | 1.20 (0.99, 1.45) | 0.066 | 1.20 (0.96, 1.50) | 0.110 |
| 45 – 54 | 2.85 (1.29, 6.31) | **0.010** | 2.45 (1.05, 5.71) | **0.037** | 1.81 (1.46, 2.24) | **<0.001** | 1.84 (1.43, 2.35) | **<0.001** |
| 55 – 64 | 2.72 (1.05, 7.07) | **0.040** | 1.71 (0.57, 5.18) | 0.340 | 1.79 (1.37, 2.34) | **<0.001** | 1.90 (1.42, 2.55) | **<0.001** |
| 65+ | 1.42 (0.31, 6.47) | 0.647 | 1.07 (0.23, 5.10) | 0.930 | 2.06 (1.48, 2.86) | **<0.001** | 2.16 (1.51, 3.08) | **<0.001** |
| **Sex** (n=6371) |  |  |  |  |  |  |  |  |
| Female | Ref |  | Ref |  | Ref |  | Ref |  |
| Male | 2.94 (1.89, 4.58) | **<0.001** | 2.95 (1.76, 4.95) | **<0.001** | 1.12 (0.98, 1.28) | 0.101 | 0.99 (0.85, 1.15) | 0.877 |
| **Mobility** (n=1499) |  |  |  |  |  |  |  |  |
| Class 1 – Higher mobility | 1.20 (0.45, 3.22) | 0.713 |  |  | 0.84 (0.64, 1.10) | 0.210 |  |  |
| Class 2 – Lower mobility | Ref |  |  |  | Ref |  |  |  |
| **Mode** (n=1499)<sup>a</sup> |  |  |  |  |  |  |  |  |
| Walk | Ref |  |  |  | Ref |  |  |  |
| Bus/taxi | 0.59 (0.16, 2.10) | 0.417 |  |  | 0.81 (0.59, 1.11) | 0.193 |  |  |
| Boda boda | 2.81 (0.35, 22.31) | 0.329 |  |  | 1.65 (0.73, 3.75) | 0.231 |  |  |
| Other | 1 |  |  |  | 0.79 (0.18, 3.57) | 0.761 |  |  |
| **Comm. distance** (n=1504) |  |  |  |  |  |  |  |  |
| ≤ 1 km | Ref |  |  |  | Ref |  |  |  |
| 1-2 | 0.17 (0.02, 1.34) | 0.092 |  |  | 0.93 (0.65, 1.32) | 0.668 |  |  |
| 2-5 | 0.89 (0.29, 2.75) | 0.843 |  |  | 0.88 (0.61, 1.26) | 0.473 |  |  |
| 5-10 | 0.33 (0.04, 2.63) | 0.294 |  |  | 0.78 (0.49, 1.24) | 0.294 |  |  |
| >10 | 0.51 (0.06, 4.14) | 0.531 |  |  | 0.68 (0.39, 1.22) | 0.196 |  |  |
| **Commute time** (n=1504) |  |  |  |  |  |  |  |  |
| ≤ 0.5 hours | Ref |  |  |  | Ref |  |  |  |
| 0.5 – 1 | 1.88 (0.42, 8.50) | 0.407 |  |  | 0.77 (0.54, 1.08) | 0.130 |  |  |
| 1 – 2 | 0.55 (0.05, 6.12) | 0.628 |  |  | 0.75 (0.48, 1.17) | 0.209 |  |  |
| > 2 | 1.99 (0.18, 22.31) | 0.576 |  |  | 1.08 (0.57, 2.04) | 0.822 |  |  |
| **Any nights (≥1) away from home in past month** (n=6346) |  |  |  |  |  |  |  |  |
| No | Ref |  |  |  | Ref |  | Ref |  |
| Yes | 1.96 (0.47, 8.11) | 0.354 |  |  | 1.80 (1.09, 2.94) | **0.020** | 1.66 (1.00, 2.74) | 0.050 |
| **Any change in residence in the past 5 years** |  |  |  |  |  |  |  |  |
| (n=6349) |  |  |  |  |  |  |  |  |
| No | Ref |  |  |  | Ref |  |  |  |
| Yes | 0.79 (0.32, 1.96) | 0.612 |  |  | 0.95 (0.74, 1.22) | 0.681 |  |  |
| **Reported Activity Locations** |  |  |  |  |  |  |  |  |
| (n=5747) |  |  |  |  |  |  |  |  |
| Home | 0.95 (0.60, 1.53) | 0.846 | 1.10 (0.67, 1.79) | 0.717 | 0.80 (0.70, 0.92) | **0.002** | 0.83 (0.72, 0.95) | **0.010** |
| Work | 0.86 (0.53, 1.39) | 0.534 | 0.54 (0.30, 0.96) | **0.036** | 1.05 (0.91, 1.21) | 0.486 | 0.93 (0.80, 1.09) | 0.393 |
| School | 0.16 (0.02, 1.15) | 0.068 | 0.18 (0.02, 1.36) | 0.096 | 0.86 (0.66, 1.11) | 0.251 | 1.01 (0.75, 1.37) | 0.933 |
| Market | 0.81 (0.47, 1.38) | 0.433 | 1.01 (0.57, 1.79) | 0.981 | 0.92 (0.79, 1.07) | 0.287 | 1.01 (0.85, 1.20) | 0.951 |
| Social | 2.15 (1.15, 4.02) | **0.017** | 1.96 (1.03, 3.74) | **0.042** | 1.43 (1.14, 1.79) | **0.002** | 1.44 (1.13, 1.82) | **0.003** |
| Church | 0.61 (0.37, 1.00) | 0.050 | 0.86 (0.51, 1.45) | 0.560 | 0.99 (0.86, 1.13) | 0.851 | 0.99 (0.85, 1.16) | 0.942 |

<sup>a</sup> Only a smaller subset responded to the work commute questions.
<sup>b</sup> Mobility variables were not significant in the bivariate analysis and were excluded in the adjusted analyses (n=5736).
<sup>c</sup> Mobility variables with the exception of overnight trips were not significant in the bivariate analysis and were excluded in the adjusted analyses (n=5733).

Regarding our secondary outcome, older age groups were more likely to report any of the four TB screening symptoms: age group 45 – 54 years (OR 1.83, 95%CI 1.43, 2.34), 55 – 64 years (OR 1.89, 95%CI 1.41, 2.54), and 65+ years (OR 2.14, 95%CI 1.50, 3.05). Reporting a social activity location was also significantly associated with a positive symptom screen (OR 1.45, 95%CI 1.15, 1.83) in adjusted analyses. Spending any nights away from one’s primary residence was significantly associated with positive symptom screen in the bivariate analysis but not when controlling for age and sex in the adjusted analysis (p-value=0.050).

In a subgroup analysis, restricted to those participants who reported social activity locations (n=488), only bars and other drinking venues were associated with a higher risk of TB (OR 4.51, 95%CI 1.58 – 12.90), even adjusting for age and sex. No statistically significant association was found for other reported social activity locations, including community venues, gaming halls, playgrounds, football grounds, social gatherings, or being with friends.

Together, these data point towards specific risk groups (i.e. men and older age groups) with higher risk of TB, though men were less likely to report symptoms. We also noted the importance of social activity locations, and in particular bars, as the only category of activity locations that was significantly associated with higher risk of TB, with odds nearly twice as high for those reporting this type of activity location compared to controls.

## Discussion

We compared the mobility patterns and activity locations of people with bacteriologically-confirmed TB and community controls across nine neighborhoods in Nairobi, Kenya. While mobility patterns differed significantly by sex and across age groups and between neighborhoods, we did not find a significant association between the specific measures of geographic mobility captured in this study, nor the constructed latent class consisting of “mobile” individuals, with respect to greater odds of active TB diagnosis. However, we found a strong positive association between reported ‘social’ activity locations and TB. Adjusting for age and sex, individuals reporting a ‘social’ activity location, for example bars or social gatherings, had almost twice the odds of having TB. Reporting other activity locations, such as home, school, market, and church, was not significantly associated with greater odds of TB diagnosis. Taken together, our results provide evidence for a behavioral risk factor for TB that is not related to work commute or mobility but instead associated with reporting time spent at ‘social’ activity locations.

Optimizing ACF for different settings requires knowledge of groups at higher risk for TB and screening locations to target case-finding efforts, particularly since large-scale ACF trials have shown mixed effectiveness in reducing population-level TB prevalence.^16,27^ Based on our findings, prior studies and global epidemiology, men are at increased risk of TB disease and should be prioritized for screening. While the reasons for increased TB risk among men have not been elucidated, studies have found that men are less likely to report TB-related symptoms, less likely to access TB testing and diagnosis, and less likely to complete treatment.^28^ Our finding of substantial heterogeneity of activity locations between different neighborhoods in Nairobi suggests opportunities to identify targeted screening locations. This variability, in both activity locations as well as mobility patterns, should be captured and modeled in future studies to incorporate local differences in activity between neighborhoods, including the risk of possible “importation” or “exportation” of TB between geographic areas.

Neighborhoods vary in their demographics, socio-economic profile, degree of inequality, patterns of mobility, and activity locations —all factors that may influence both transmission of TB and possible case finding strategies.

Identification of ‘social’ activity locations, in this study, as being locations more likely to be frequented by individuals with TB provides potential venues for ACF activities, both in Nairobi and possibly other high-burden urban settings. Rather than a costlier broad approach, a spatially-targeted approach would deploy ACF activities to specific locations known or suspected of being TB transmission ‘hotspots’.^17,29^ Other studies have pointed to healthcare facilities and markets as important venues for likely TB transmission.^19,20,30^ We found reporting ‘social’ activity locations, particularly bars and other drinking venues, to be highly associated with TB diagnosis. Investigating the extent of this association and identifying particular social venues can help to inform national and local TB programs on where to screen for TB or implement environmental modifications, such as improved ventilation, to reduce the risk of TB transmission. Implementation studies will be critical for understanding the acceptability of TB screening in community venues. Combining granular activity locations data with results from a recent prevalence survey or modelled TB incidence estimates^31^ can provide a data-driven approach to spatial targeting of ACF in a community.

Our study has several limitations. One major limitation is the absence of geospatial data and detailed information about the duration and nature of time spent at various activity locations. Optimally, activity spaces data would be captured from a case-control study with in-depth interviews and geo-tracking of individuals both with and without TB. Participation in the work commute survey was limited to only those who provided a site or location for employment, leading to selection bias when collecting mobility data. Summary statistics of mobility characteristics were not representative of the total population, with responses totaling less than a quarter of all participants. Moreover, daily commute and mobility are not constrained to only travel to and from work. Individuals may move about outside of their homes for any number of reasons, work or non-work related. Capturing data on all types of movement, not only work commute, can give a more comprehensive picture of an individual’s mobility patterns and associated risks and exposures. Additionally, selection bias is possible within the overall prevalence survey, as we could only enroll individuals that were present at home during the census and were able to present to the mobile field site for symptom screen and chest X-ray. Second, our study was limited by the unstructured responses allowed for reporting activity locations hindering our ability to extract detailed geographic information on specific physical venues. Abstract locations, for example “Work place” or “market”, could not be geo-located, thus our analysis was constrained to broad categories of activity locations, such as home, work, social, market, etc. It is also likely for some degree of bias to have been introduced via our post-hoc categorization of activity locations. Additionally, specifying a more explicit time frame for activity locations, such as in the past 30 days prior to diagnosis, may help with both recall and point towards sites of possible recent transmission. Third, our analysis focused on the primary outcome of bacteriologically-confirmed TB diagnosis, either by sputum culture and/or Xpert Ultra. With only a small number of positive cases found overall in the prevalence survey (∼1%), and even fewer positive cases that completed the work commute questionnaire (n=73), we were very likely to have been underpowered to identify significant differences in exposures between TB cases and controls. Finally, enrollment in the parent study occurred as Kenya was exiting COVID-19 pandemic-era mandates. It is unclear to what extent these mandates restricted movement or altered habits compared to pre- or post-pandemic.

Future work to investigate the association of mobility and activity locations with TB, particularly with the goal of informing targeted active case finding activities, may benefit from a more quantitative and detailed approach at capturing dynamic patterns of movement. Studies utilizing GPS tracking, for example, have reported on travel patterns and activity spaces of patients with MDR-TB,^32^ pregnant women,^33^ residents of malaria-endemic regions,^34^ and young adults with high risk of HIV acquisition.^35^ Comprehensive evaluation of mobility, however, will likely require not only GPS tracking but also qualitative interviews in a mixed methods approach to truly understand the varied contexts and attributes relating to travel. Travel diaries, GPS loggers, and focused interviews in tandem will likely bring out more useful and relevant information with regards to human movement and exposures. Understanding heterogeneous patterns of mobility and activity in various contexts, settings, and environments and among different sub-populations and across regions may help to identify those groups with the highest risk of TB exposure and where to find them.

In summary, mobility patterns and activity locations offer distinct perspectives on human behavior not traditionally captured in standard epidemiological studies. Investigating these geographic characteristics, at both the individual and population level, can not only offer insights on risks and exposures for TB transmission but also provide practical insights for optimizing targeted active case finding strategies.

## Data Availability

All data produced in the present study are available upon reasonable request to the authors.

## Acknowledgements

We would like to thank the study participants, the community health workers, the KEMRI CRDR Nairobi staff, and the study team.

