## Supplement for "Mobility patterns, activity locations, and tuberculosis in Nairobi, Kenya"

**Table S1. Features of geographic mobility**

| **Variable** | **Higher mobility** | **Lower mobility** |
| --- | --- | --- |
| Mode of transportation (working commute) | Bus, taxi, or other vehicles | Walk |
| Commute distance  *Sensitivity analysis*: *different thresholds* | > 2km | <= 2km |
| Commute time | > 0.5 hour | <= 0.5 hour |
| Transit hub – Do you use a transit hub (e.g. bus station or taxi rank) | Yes | No |
| Travel away from home in past month, # of nights | 1 or more nights | 0 nights |
| Residence change – Have you changed your residence at any point the last 5 years | Yes | No |

**Table S2. LCA model selection: # classes, degrees of freedom, BIC, AIC**

| **Number of Latent Classes** | **Degrees of freedom** | **Bayesian Information Criterion** | **Akaike’s Information Criterion** |
| --- | --- | --- | --- |
| 1 | 6 | 7174.1 | 7142.2 |
| 2 | 13 | 6446.3 | 6377.2 |
| 3 | 20 | No convergence |  |
| 4 | 27 |  |  |

**Table S3. Estimated marginal probabilities for individual features in each latent class of mobility**

|  | **Class 1**  **(Higher mobility)**  **Mean (95%CI)** | **Class 2**  **(Lower mobility)**  **Mean (95%CI)** | **Difference**  **(Class 1 – Class 2)** |
| --- | --- | --- | --- |
| Commute distance > 2km | 0.92 (0.86 – 0.96) | 0.06 (0.03 – 0.12) | 0.86 |
| Bus/taxi or other mode of transportation (not walking) | 0.70 (0.64 – 0.75) | 0.03 (0.01 – 0.07) | 0.67 |
| Commute time > 0.5 hours | 0.97 (0.95 – 0.98) | 0.70 (0.66 – 0.73) | 0.27 |
| Spends time at transit hub | 0.09 (0.07 – 0.12) | 0.04 (0.03 – 0.06) | 0.05 |
| Traveled away from home >= 1 night in last month | 0.03 (0.02 – 0.05) | 0.02 (0.01 – 0.03) | 0.01 |
| Changed residence in last 5 years | 0.06 (0.04 – 0.08) | 0.08 (0.06 – 0.10) | -0.02 |

**Figure S1. Marginal probabilities for mobility features in each latent class of mobility**

|  | Class 1: Higher Mobility | Class 2: Lower Mobility |
| --- | --- | --- |
| Bus/taxi or other mode of transportation (not walking) | 0.695942 | 0.0306795 |
| Commute distance >= 2km | 0.919573 | 0.0625898 |
| Commute time >= 0.5 hrs | 0.969845 | 0.696718 |
| Spends time at transit hub | 0.0933852 | 0.0421119 |
| Traveled away from home >= 1 night | 0.0296733 | 0.0151676 |
| Changed residence in last 5 years | 0.056489 | 0.0780465 |
